# Genetic evidence prioritizes circulating proteins for heart failure beyond shared BMI-related genetic liability

**DOI:** 10.64898/2026.06.19.26356109

**Authors:** Chen-Yang Su, Tianyuan Lu

**Author notes:** **Correspondence:** Chen-Yang Su, PhD; Address: 1007E-610 Walnut St, Madison, WI 53726, USA; Tianyuan Lu, PhD; Address: 1007E-610 Walnut St, Madison, WI 53726, USA.

## Abstract

**Background:** Heart failure (HF) and body mass index (BMI) share substantial genetic architecture, which may lead genetically informed target discovery to preferentially identify adiposity-related pathways. We sought to identify circulating proteins associated with HF beyond this shared genetic component.

**Methods:** We applied GWAS-by-subtraction to overall HF, nonischemic HF, and nonischemic HF with reduced or preserved ejection fraction to derive BMI-related and BMI-subtracted HF components. We then performed proteome-wide *cis*-pQTL Mendelian randomization and colocalization using four independent proteomic cohorts, followed by tissue-specific eQTL colocalization, cardiac transcriptomic annotation, and druggability assessment.

**Results:** Compared with the original HF phenotypes, the BMI-subtracted components showed attenuated genetic correlations with BMI (0.045–0.147) while retaining 28 independent loci for overall HF and nine for nonischemic HF. Across 19,930 protein-HF tests, 11 associations involving nine proteins were prioritized by the Mendelian randomization and colocalization analyses. For example, a 1-SD increase in genetically predicted CELSR2 abundance was associated with lower overall HF risk (odds ratio, 0.96 [95% CI, 0.94–0.98]; *P*=8.6×10^-7^), whereas a 1-SD increase in genetically predicted CSF3 abundance was associated with higher nonischemic HF risk (odds ratio, 1.32 [95% CI, 1.18–1.48]; *P*=2.0×10^-6^). CELSR2 and TMEM106B colocalized with *cis*-eQTLs in failing left ventricular myocardium, and DAG1 showed cardiomyocyte enrichment with concordant downregulation in failing hearts.

**Conclusions:** We identified nine circulating proteins associated with HF beyond the genetic component shared with BMI. These findings extend the range of genetically supported pathways implicated in HF and nominate candidate proteins for further mechanistic and therapeutic investigation.

**Clinical Perspective:** *What Is New?:*

- GWAS-by-subtraction separated HF genetic susceptibility into BMI-related and BMI-subtracted components, allowing proteome-wide target discovery to focus on HF associations beyond the genetic component shared with BMI.
- Proteome-wide Mendelian randomization and colocalization prioritized nine circulating proteins.
- Integration with cardiac molecular data provided additional support for selected proteins, including failing-left-ventricular eQTL colocalization for CELSR2 and TMEM106B and cardiomyocyte expression with concordant myocardial downregulation for DAG1.

*What Are the Clinical Implications?:*

- Accounting for shared BMI-related genetic liability may extend human genetics-guided target discovery to HF pathways that could be overlooked when conventional HF GWAS are used as the disease outcome.
- The findings illustrate that therapeutic interpretation must consider the genetically supported direction of effect. The LPA association was consistent with Lp(a) lowering, whereas the CSF3 and RSPO3 results did not support straightforward repurposing of existing pharmacological strategies.
- The prioritized proteins provide candidates for further functional and therapeutic investigation, but replication and experimental validation are needed before their clinical relevance can be established.

## Introduction

Despite major advances in pharmacological and device-based treatment, heart failure (HF) remains a leading cause of morbidity, mortality, and hospitalization^1,2^. HF is a heterogeneous clinical syndrome that represents the common manifestation of multiple upstream pathophysiological processes^3–5^. This biological heterogeneity presents a major challenge for therapeutic development, as distinct molecular mechanisms may contribute to HF susceptibility across individuals and disease subtypes. Identifying biological pathways that directly influence HF susceptibility remains an important priority for improving prevention and treatment.

Human genetics offers an opportunity for therapeutic target discovery^6^. Mendelian randomization (MR) uses genetic variants associated with a potentially modifiable exposure as instruments to assess its association with disease risk, reducing susceptibility to confounding and reverse causation under its assumptions^7^. In circulating proteome-wide MR, *cis*-acting protein quantitative trait loci (*cis*-pQTL) can be used to evaluate whether genetically predicted circulating protein abundance is associated with HF^8–11^. When combined with genetic colocalization analyses, which evaluate whether protein and disease associations are attributable to a shared causal variant, these approaches can provide evidence linking inherited genetic variation to potentially actionable therapeutic targets^8,12^.

A central challenge, however, is that MR target discovery depends on the genetic architecture of the disease outcome used in the analysis. Body mass index (BMI) is among the strongest risk factors for HF^13–15^, and large HF genome-wide association studies (GWAS) show substantial genetic overlap with BMI^4,5^. Consequently, proteome-wide MR using conventional HF GWAS may preferentially identify proteins whose associations with HF reflect adiposity or adiposity-related pathways. Such pathways are clinically relevant, but they may not capture the full spectrum of biological processes contributing to HF susceptibility. Disentangling obesity-related mechanisms from other contributors to HF may therefore broaden the range of genetically supported therapeutic targets and provide insight into pathways that operate beyond adiposity-mediated disease risk.

GWAS-by-subtraction offers a way to address this limitation by separating the genetic component shared between correlated traits from variation more specific to each trait^16,17^. We applied GWAS-by-subtraction to overall HF, nonischemic HF (niHF), nonischemic HF with reduced ejection fraction (niHFrEF), and nonischemic HF with preserved ejection fraction (niHFpEF) to derive BMI-related and BMI-subtracted HF components. We then performed proteome-wide *cis*-pQTL MR and colocalization against the BMI-subtracted components using four independent proteomic cohorts. Finally, we integrated tissue and failing-heart eQTL colocalization, cardiac cell-type expression, myocardial differential expression, cross-cohort evidence, and druggability to prioritize circulating proteins associated with HF genetic susceptibility beyond the component shared with BMI.

## Methods

### Heart failure and BMI GWAS summary statistics

We used heart failure summary statistics from individuals of European ancestry from the HERMES Consortium 2.0 case-control GWAS meta-analysis for four phenotypes: overall heart failure (HF; 139,533 cases), nonischemic HF (niHF) excluding coronary, valvular, and congenital disease (42,081 cases), nonischemic HF with reduced ejection fraction (niHFrEF; 4,975 cases), and nonischemic HF with preserved ejection fraction (niHFpEF; 3,590 cases)^5^. The GWAS included up to approximately 1.7 million individuals. BMI summary statistics were obtained from the GIANT and UK Biobank meta-analysis reported by Yengo et al. (N = 681,275)^18^ (**Supplementary Table 1**).

### GWAS-by-subtraction

We used Genomic Structural Equation Modeling (GenomicSEM) to decompose each HF GWAS with respect to BMI using a GWAS-by-subtraction approach^16,17^. Model details have been previously described^19,20^. Summary statistics were formatted and restricted to HapMap3 variants. Genetic covariance between BMI and each HF phenotype was estimated by LD-score regression using European LD scores^21,22^. For each HF phenotype, we fit a genomic structural equation model comprising two latent factors, with a BMI-related factor loading on both BMI and HF and a BMI-subtracted factor loading on HF only. SNP effects on these latent factors were estimated throughout the genome. This procedure generated two SNP-level GWAS per HF phenotype, corresponding to the BMI-related HF component and the BMI-subtracted HF component.

### Validation of the GWAS-by-subtraction decomposition

We verified the validity of GWAS-by-subtraction by estimating the genetic correlation of each BMI-related and BMI-subtracted factor with BMI using bivariate LD-score regression. Summary statistics were restricted to high-quality HapMap3 variants outside the MHC region and analysed using European ancestry LD scores. Successful decomposition was defined by strong genetic correlation between the BMI-related factor and BMI and minimal residual genetic correlation between the BMI-subtracted factor and BMI. LD-score-regression intercept was used to assess residual confounding. Genome-wide-significant variants were defined as autosomal variants with *P* < 5 × 10^-8^. Independent loci were identified by greedy 1-Mb distance-based clumping of significant variants (lowest *P* variant as lead, all variants within 1 Mb on the same chromosome assigned to that locus).

### *cis*-pQTL instruments and proteome-wide MR

We leveraged four European ancestry affinity-proteomic cohorts: ARIC, deCODE and Fenland measured on the SomaScan v4 aptamer-based platform, and UKB-PPP measured on the antibody-based Olink Explore 3072 platform^23–26^. Variants were LD clumped (*r^2^* < 0.001, 1 Mb window, and *P* < 5 × 10^-8^) using the 1000 Genomes European reference panel^27^. *Cis*-pQTL were defined as genome-wide significant variants (*P* < 5 × 10^-8^) located within 1 Mb of the transcription start site of the corresponding protein-coding gene on genome build GRCh38.

Protein abundance was normalized within each study; therefore, MR effect estimates are expressed per standard deviation increase in genetically predicted protein abundance. Exposure and outcome summary statistics were harmonized to the same effect allele. Palindromic variants with ambiguous allele frequencies (MAF > 0.42) were excluded. All analyses were restricted to European ancestry.

For each protein-HF pair, we performed two-sample MR against the BMI-subtracted HF GWAS using the TwoSampleMR package. We used the Wald ratio for proteins instrumented by a single *cis*-pQTL and inverse-variance weighting for proteins instrumented by two or more *cis*-pQTLs^28^. Effects are reported as odds ratios per standard deviation increase in genetically predicted protein abundance.

For each instrument, F-statistics were calculated as (β/SE)^2^ using the *cis*-pQTL effect estimate and standard error^29^. Cohort-level instrument strength was summarized using the regression F-statistic computed as F = R^2^(N − k − 1)/[k(1 – R^2^)], where R^2^ is the proportion of protein abundance variance explained, N is the *cis*-pQTL sample size, and *k* is the number of instruments. All retained instruments had F > 10. For proteins with two or more instruments, heterogeneity was assessed using Cochran’s Q from the inverse-variance-weighted model, with *I^2^* calculated as max{0, (Q − df)/Q}. MR-Egger^30^, weighted-median^31^, and weighted-mode estimates^32^, together with the MR-Egger intercept, were computed for proteins with three or more instruments. Steiger directionality tests were used to compare the variance explained in protein abundance with the variance explained in HF^33^. Within each cohort, multiple testing was controlled using the Benjamini-Hochberg procedure^34^, with an FDR q value <0.05 considered significant. We additionally applied a study-wide Bonferroni threshold of *P* <0.05/19,930 = 2.5×10^-6^ across all protein-HF tests. Reporting followed the STROBE-MR guidelines (**Supplementary Note 1**).

### Colocalization of pQTL and HF signals

For each MR-significant association, we tested whether the *cis*-pQTL and BMI-subtracted HF association signals colocalized using PWCoCo^8^ and SharePro^12^. PWCoCo performs pairwise conditional and colocalization analysis, whereas SharePro allows for multiple causal variants and estimates the probability of a shared signal. Associations were considered colocalized if the PWCoCo posterior probability for a shared causal variant, PP.H4, or the SharePro shared-signal probability was ≥ 0.8. For each protein, we reported the maximum colocalization probability across PWCoCo and SharePro.

### Tissue and cardiac eQTL colocalization

To assess whether genetically predicted protein abundance and gene expression were influenced by shared causal variants, we performed colocalization analyses between *cis*-pQTL and *cis*-eQTL from Genotype-Tissue Expression (GTEx) v8^35^ across 13 tissues and from the TOPCHeF cohort^36^ of failing left ventricular myocardium, which comprised approximately 516 participants with RNA sequencing and whole-genome sequencing data. Colocalization was performed using SharePro and the 1000 Genomes European LD reference panel. Evidence of colocalization was defined as PP.H4 ≥ 0.8. For each protein-gene pair, the maximum colocalization probability was retained across genetic instruments, proteomic cohorts, and, for GTEx, the 13 evaluated tissues.

### Cardiac cell-type and myocardial-expression annotation

To characterize the cardiac cellular distribution and heart failure-related regulation of the prioritized proteins, we annotated their corresponding genes using ReHeaT2, an integrated resource comprising 25 bulk-tissue and single-nucleus transcriptomic studies of heart failure^37^. Consensus markers derived from the single-nucleus datasets were used to identify the cardiac cell type or cell types in which each gene was expressed. Consensus differential expression estimates from bulk myocardial tissue were used to determine whether each gene was upregulated or downregulated in failing relative to nonfailing myocardium. The direction of myocardial differential expression was subsequently compared with the direction of the MR association to classify the evidence as directionally concordant or discordant.

### Druggability annotation

We classified proteins according to the druggable genome tiers defined by Finan et al.^38^, which rank proteins according to their tractability for pharmacologic targeting: Finan Tier 1, targets of approved drugs or drugs in clinical development; Finan Tier 2, proteins with high sequence similarity to Finan Tier 1 targets or known to bind drug-like small molecules; and Finan Tier 3, secreted or extracellular proteins and members of major druggable gene families, including G-protein-coupled receptors, kinases, ion channels, and nuclear receptors, not assigned to Finan Tier 1 or Finan Tier 2. Finan Tier 3 was further subdivided into Finan Tier 3A, comprising proteins with greater similarity to established drug targets, and Finan Tier 3B, comprising the remaining proteins. To distinguish these categories from the integrated prioritization tiers used in this study, we refer to them throughout as “Finan T1” through “Finan T3B.” We additionally queried the Open Targets Platform to identify drugs known to target each protein and recorded the highest reported phase of clinical development^39^. Drug annotations were interpreted in relation to the direction of the MR estimate to determine whether higher or lower protein abundance represented the direction associated with reduced heart failure risk.

### Integrated prioritization

We integrated evidence from *cis*-pQTL MR, plasma *cis*-pQTL colocalization with heart failure, tissue and failing-heart *cis*-eQTL colocalization, myocardial expression analyses, cross-cohort replication, and druggability into a unified target prioritization framework. Tier 1 included proteins with MR associations surpassing the study-wide Bonferroni threshold (*P* < 2.5 × 10^-6^) and strong plasma *cis*-pQTL colocalization, defined as a PWCoCo or SharePro PP.H4 ≥ 0.8. Tier 2 included proteins that met the FDR threshold, did not reach the study-wide Bonferroni threshold, and showed strong plasma *cis*-pQTL colocalization together with at least one additional supporting feature. Supporting features included *cis*-eQTL colocalization in failing left ventricular myocardium from TOPCHeF or in a GTEx tissue (PP.H4 ≥ 0.8), directional concordance between the MR association and differential myocardial expression, MR support in more than one proteomic cohort, or the availability of a clinical-stage drug targeting the protein. Tier 3 included the remaining proteins that met the FDR threshold and showed strong plasma *cis*-pQTL colocalization but lacked an additional supporting feature.

### Data availability

Heart failure summary statistics are publicly available; pQTL and eQTL summary statistics are available from the respective source studies.

## Results

### GWAS-by-subtraction identifies BMI-subtracted genetic components of HF

We applied GWAS-by-subtraction to four HF phenotypes (overall heart failure [HF], nonischemic HF [niHF], nonischemic HF with reduced ejection fraction [niHFrEF], and nonischemic HF with preserved ejection fraction [niHFpEF]) and BMI. Each HF phenotype was modeled in conjunction with BMI to derive a BMI-related latent factor, which loaded on both HF and BMI, and a BMI-subtracted factor, which loaded only on HF. Genetic correlation analyses supported the intended decomposition (**Figure 1a**; **Supplementary Table 2**). The original HF phenotypes showed moderate genetic correlations with BMI (*r_g_* = 0.23 for niHFrEF to *r_g_* = 0.58 for niHFpEF), reflecting the shared genetic component targeted by the decomposition. In contrast, the BMI-subtracted HF factors showed substantially attenuated genetic correlations with BMI (*r_g_* =0.045 for niHFrEF to *r_g_* =0.147 for overall HF; **Supplementary Table 2** and **Figure 1b**).

**Figure 1.**
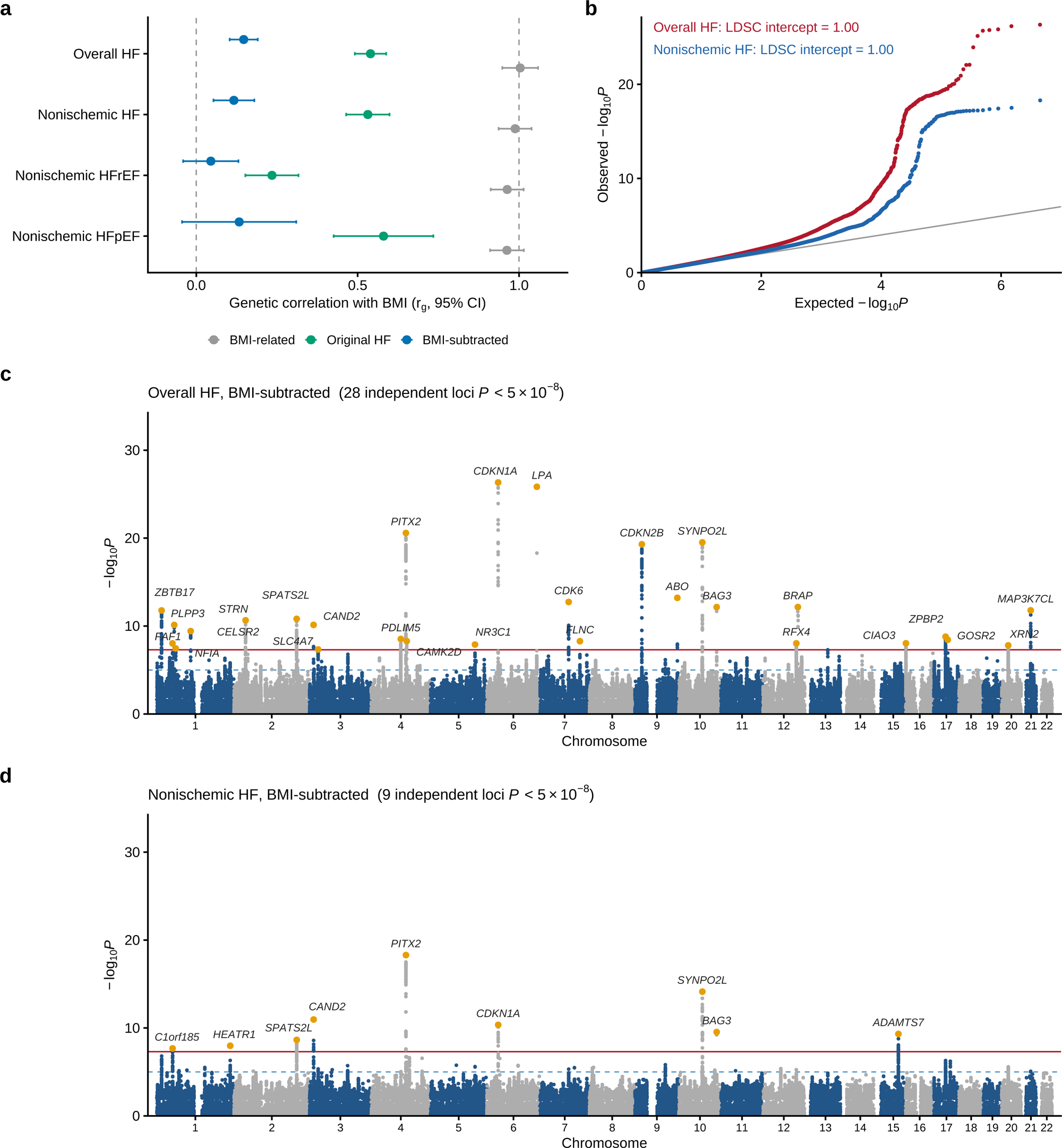
GWAS-by-subtraction of heart failure on BMI. (a) Genetic correlation with BMI, estimated by LD-score regression, for each original HF trait (green), the BMI-related latent factor (gray), and the BMI-subtracted latent factor (blue). Points show estimates and horizontal bars show 95% confidence intervals; dashed vertical lines mark genetic correlations of 0 and 1. Results are shown for overall HF, nonischemic HF, nonischemic HFrEF and nonischemic HFpEF. (b) Quantile-quantile plots of the BMI-subtracted overall HF (red) and nonischemic HF (blue) GWAS. The LD-score regression intercept is shown for each. (c, d) Manhattan plots of the BMI-subtracted overall HF (c) and nonischemic HF (d) GWAS. The red horizontal line marks genome-wide significance (*P* = 5 × 10^-8^), and the blue dashed line marks the suggestive threshold (*P* = 1 × 10^-5^); alternating colors distinguish chromosomes. Lead variants at genome-wide significant loci are highlighted in orange and labeled with the nearest protein-coding gene based on RefGene hg19 annotation. Leader lines connect each lead variant to its label, and the full locus list is provided in **Supplementary Table 3**.

The BMI-subtracted factors retained genome-wide significant association signals for the two best-powered HF phenotypes (**Figure 1c** and **Figure 1d**). The BMI-subtracted overall HF GWAS identified 28 genome-wide significant independent loci, whereas the niHF GWAS identified 9 loci (**Supplementary Table 3**). Leading loci for overall HF included established cardiovascular regions, including the LPA locus on chromosome 6 (rs10455872, *P*=1.5×10^-26^) and the chromosome 1p13 CELSR2 lipid locus (rs12740374, *P*=3.8×10^-10^). By contrast, the lower-powered ejection-fraction subtypes yielded fewer associations. niHFrEF identified 7 genome-wide significant indepedent loci, whereas niHFpEF, which included only 3,590 cases, identified no genome-wide significant variants (**Supplementary Table 3**; **Supplementary Figure 1**).

### Proteome-wide *cis*-pQTL MR and colocalization prioritize nine proteins

We tested *cis*-pQTL instruments from 4 proteomic cohorts (ARIC, deCODE, and Fenland using SomaScan and UK Biobank Pharma Proteomics Project [UKB-PPP] using Olink) against the four BMI-subtracted HF phenotypes, comprising 19,930 protein-HF tests (**Supplementary Table 4**). Given the limited power of the ejection-fraction subtype GWAS, protein prioritization was based on BMI-subtracted overall HF and nonischemic HF; all associations meeting the prioritization criteria arose from these two phenotypes. Prioritization required both a within-cohort false discovery rate (FDR)-significant MR association and evidence that the *cis*-pQTL and HF association shared an underlying genetic signal, as assessed using PWCoCo or SharePro (PP.H4 ≥0.8). Across the four proteomic cohorts, 11 associations, corresponding to nine unique proteins, met both criteria (**Figure 2**; **Supplementary Table 5**). An additional six associations met the within-cohort FDR threshold but lacked evidence of colocalization and therefore were not prioritized. These included FURIN and PLAU, which were associated with BMI-subtracted HF in MR analyses but did not colocalize with the corresponding HF association signal (**Supplementary Table 6**). We verified that the strength of all instruments was sufficient (F > 10; **Supplementary Tables 7** and **8**).

**Figure 2.**
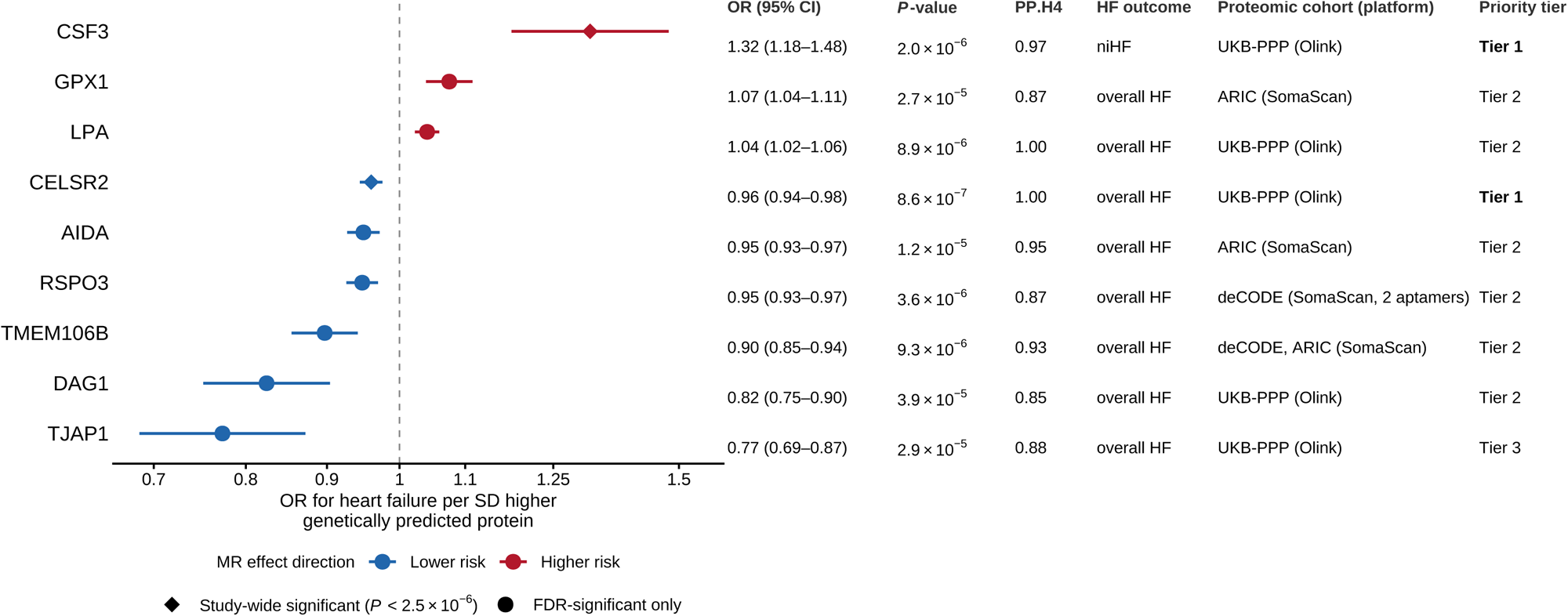
*cis*-pQTL Mendelian randomization and colocalization prioritize nine proteins for BMI-subtracted heart failure. Forest plot of the odds ratio for HF per SD increase in genetically predicted protein abundance. Points show odds ratios, horizontal lines show 95% confidence intervals, and the dashed vertical line marks OR = 1. Points are colored according to the direction of the MR association, with blue indicating lower HF risk and red indicating higher HF risk. Diamonds mark proteins that reached the study-wide Bonferroni threshold (*P* < 2.5 × 10^-6^), whereas circles identify proteins that met the within-cohort false discovery rate threshold but not the study-wide Bonferroni threshold. The right panel reports the OR and 95% CI, MR *P*-value, the *cis*-pQTL–HF colocalization probability (taken as the larger of the PWCoCo and SharePro probabilities), the HF outcome, proteomic cohort and assay platform, and integrated priority tier for each protein. Each protein contributes one row: RSPO3 was assayed by two deCODE SomaScan aptamers, both prioritized, and is shown once using the better-powered aptamer. Counting both RSPO3 aptamers and both TMEM106B cohorts, these nine proteins correspond to 11 prioritized associations.

Of the nine prioritized proteins, two met the study-wide Bonferroni significance threshold (*P* <0.05/19,930 = 2.5×10^-6^). Higher genetically predicted CELSR2 abundance was associated with lower risk of overall HF (OR per SD, 0.96 [95% CI, 0.94–0.98]; *P* = 8.6×10^-7^; UKB-PPP), with strong evidence of colocalization (PP.H4 = 1.00). Higher genetically predicted abundance of CSF3, which encodes granulocyte colony-stimulating factor, was associated with higher risk of niHF (OR, 1.32 [95% CI, 1.18–1.48]; *P* = 2.0×10^-6^; UKB-PPP), with similarly strong colocalization evidence (PP.H4 = 0.97).

The remaining 7 proteins met the within-cohort false discovery rate threshold but not the study-wide Bonferroni threshold, and all showed strong evidence of colocalization. Higher genetically predicted abundances of RSPO3 (OR, 0.95 [95% CI, 0.93–0.97]; *P* = 3.6×10^-6^; deCODE; PP.H4=0.87), TMEM106B (OR, 0.90 [95% CI, 0.85–0.94]; *P* = 9.3×10^-6^; deCODE; PP.H4=0.93), AIDA (OR, 0.95 [95% CI, 0.93–0.97]; *P* = 1.2×10^-5^; ARIC; PP.H4=0.95), DAG1 (OR, 0.82 [95% CI, 0.75–0.90]; *P* = 3.9×10^-5^; UKB-PPP; PP.H4=0.85), and TJAP1 (OR, 0.77 [95% CI, 0.69–0.87]; *P* = 2.9×10^-5^; UKB-PPP; PP.H4=0.88) were associated with lower risk of overall HF. Conversely, higher genetically predicted abundances of LPA (OR, 1.04 [95% CI, 1.02–1.06]; *P* = 8.9×10^-6^; UKB-PPP; PP.H4=1.00) and GPX1 (OR, 1.07 [95% CI, 1.04–1.11]; *P* = 2.7×10^-5^; ARIC; PP.H4=0.87) were associated with higher risk.

### Tissue eQTL colocalization provides cardiac and metabolic tissue support for selected proteins

To evaluate whether the genetic determinants of circulating protein abundance overlapped with gene regulatory signals in disease-relevant tissues, we colocalized each protein’s *cis*-pQTL with *cis*-eQTLs from 13 GTEx tissues and failing left ventricular myocardium from TOPCHeF (**Supplementary Tables 9** and **10**; **Figure 3**).

**Figure 3.**
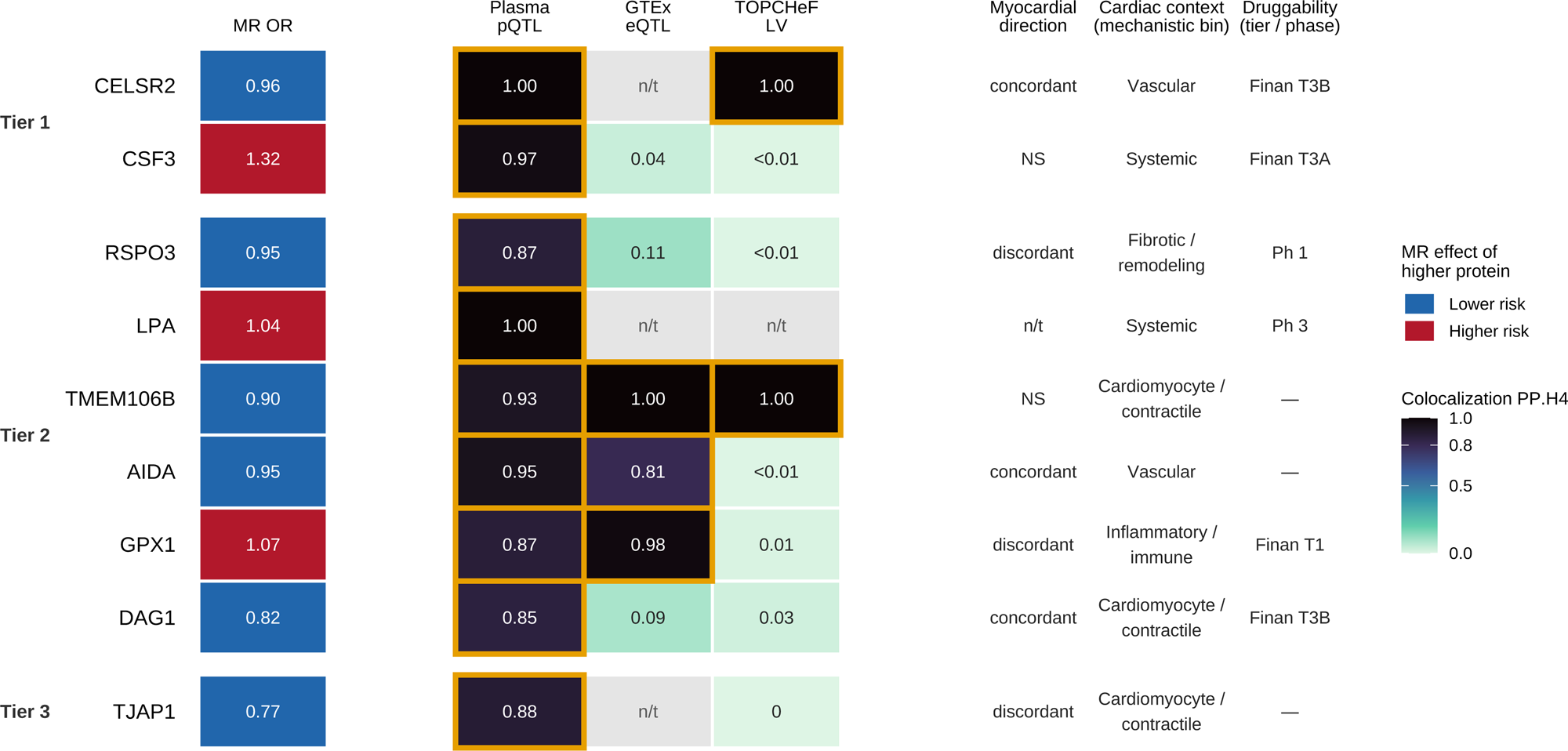
Integrated evidence supporting the prioritized proteins. Heatmap summarizing genetic, tissue-regulatory, myocardial-expression, cardiac-context, and druggability evidence for the nine prioritized proteins, grouped by integrated priority tier. The MR OR column shows the odds ratio for HF per SD increase in genetically predicted protein abundance, with blue indicating lower HF risk and red indicating higher HF risk. The three colocalization columns report the probability of a shared genetic signal between the *cis*-pQTL and, respectively, HF for the best-powered instrument, a GTEx *cis*-eQTL based on the maximum probability across 13 tissues, and a TOPCHeF *cis*-eQTL in failing left ventricular myocardium. For the plasma pQTL column the value is the larger of the PWCoCo and SharePro probabilities; the GTEx and TOPCHeF eQTL columns report the SharePro shared-signal probability. Cells with values ≥0.80 are outlined in gold. The remaining columns summarize directional concordance between the MR association and myocardial differential expression, a cardiac mechanistic bin derived from each protein’s top ReHeaT2 consensus-marker cell type (per-protein cell types in **Supplementary Table 11**), and druggability. “NS” indicates no significant myocardial differential expression, “n/t” indicates that the comparison was not testable, and “—” indicates that no corresponding druggability annotation was identified. Druggability is reported as the Finan druggability tier (Finan T1–T3B) or the highest clinical development phase (Ph) to distinguish these annotations from the integrated priority tiers.

Across GTEx tissues, *TMEM106B* showed the broadest evidence of regulatory overlap. Its *cis*-pQTL colocalized with the *TMEM106B cis*-eQTL in left ventricular myocardium, atrial appendage, and tibial artery (≥ 0.99), with additional support in whole blood (PP.H4 = 1.0), adrenal gland (PP.H4 = 1.0), and skeletal muscle (PP.H4 = 0.94). *GPX1* showed evidence of colocalization in skeletal muscle (PP.H4 = 0.98), whereas *AIDA* colocalized in adrenal gland (PP.H4 = 0.81). By contrast, *RSPO3* showed no evidence of colocalization in the GTEx tissues examined (PP.H4 = 0.11 in subcutaneous adipose tissue), and no colocalization was observed for *DAG1* or *CSF3*. *CELSR2*, *LPA*, and *TJAP1* could not be evaluated because the corresponding genes were not testable in the selected GTEx tissues.

The strongest cardiac tissue support was observed in TOPCHeF failing left ventricular myocardium. The *cis*-pQTLs for *TMEM106B* and *CELSR2* each colocalized with their corresponding cardiac *cis*-eQTL (PP.H4 = 1.00), providing convergent cardiac tissue regulatory support; no other prioritized protein showed comparable evidence (**Supplementary Table 10**).

### Cardiac cell-type expression and concordance with myocardial regulation

We annotated the nine prioritized proteins for cardiac cell-type expression and myocardial regulation using ReHeaT2, an integrated analysis of 25 bulk-tissue and single-nucleus transcriptomic HF datasets (**Supplementary Table 11**; **Supplementary Figure 2**). These proteins mapped to multiple cardiac lineages. Based on their top ReHeaT2 consensus-marker cell type, DAG1, TJAP1, and TMEM106B were cardiomyocyte markers; CELSR2 was a pericyte marker, with vascular smooth muscle as a secondary marker; AIDA was an endothelial marker; RSPO3 was a fibroblast marker; and GPX1 was a myeloid marker.

For three proteins, the direction of the MR association was concordant with the direction of differential myocardial expression in HF. DAG1 showed the clearest convergence of genetic and myocardial-expression evidence. It was identified as a cardiomyocyte marker and was markedly downregulated in failing myocardium (log fold-change, −0.21; *P* = 4.5×10^-38^), whereas higher genetically predicted DAG1 abundance was associated with lower HF risk (OR, 0.82). CELSR2 and AIDA showed similar directional concordance. Both were downregulated in failing myocardium (CELSR2 log fold change -0.11; *P* = 1.7×10^-3^; AIDA log fold change, -0.12; *P* = 7.6×10^-8^), and higher genetically predicted abundance of each was associated with lower HF risk (CELSR2 OR, 0.96; AIDA OR, 0.95).

GPX1, RSPO3, and TJAP1 showed discordant directions between the MR and myocardial-expression analyses. However, such discordance should be interpreted cautiously because circulating protein abundance and myocardial transcript expression represent distinct molecular traits, and differential expression in established HF may reflect compensatory responses, treatment effects, altered cell composition, or consequences of disease.

### Druggability and therapeutic context

We mapped the nine proteins to the druggable genome and to drug target evidence in Open Targets (**Supplementary Table 12**). LPA is targeted by two investigational Phase 3 Lp(a)-lowering therapies, the antisense oligonucleotide pelacarsen and the small interfering RNA olpasiran. Because higher genetically predicted LPA abundance was associated with increased HF risk, therapeutic Lp(a) lowering was directionally concordant with the MR estimate, providing a positive control example. RSPO3 is targeted by rosmantuzumab, an anti-RSPO3 monoclonal antibody previously evaluated in a Phase 1a/b oncology trial. However, RSPO3 inhibition was directionally discordant with the MR estimate, which associated higher genetically predicted RSPO3 abundance with lower HF risk.

GPX1 was classified as a Finan Tier 1 druggable target, although no corresponding drug was identified in Open Targets, whereas CELSR2 and DAG1 were classified as predicted biologic-druggable targets (Finan Tier 3B). CSF3 encodes granulocyte colony-stimulating factor, for which recombinant G-CSF therapies, including filgrastim and pegfilgrastim, are clinically available^40^. However, because higher genetically predicted CSF3 abundance was associated with increased nonischemic HF risk, the MR findings were directionally discordant with therapeutic CSF3 augmentation and should not be interpreted as supporting such an intervention.

### Integrated evidence-based prioritization

We integrated all evidence layers to assign each protein to an evidence-based priority tier (**Figure 3**; **Supplementary Table 13**). Tier 1 included proteins with study-wide Bonferroni-significant MR associations and strong *cis*-pQTL-HF colocalization. Tier 2 included proteins that met the within-cohort false discovery rate threshold but not the study-wide Bonferroni threshold, showed strong *cis*-pQTL-HF colocalization, and had support from at least one additional evidence layer. These supporting layers comprised cardiac or GTEx tissue eQTL colocalization, directional concordance between the MR association and differential myocardial expression, support across multiple proteomic cohorts, or an existing clinical-stage drug targeting the protein. Proteins not meeting the Tier 1 or Tier 2 criteria were assigned to Tier 3.

Accordingly, CELSR2 and CSF3 were assigned to Tier 1; TMEM106B, DAG1, RSPO3, LPA, AIDA, and GPX1 to Tier 2; and TJAP1 to Tier 3. Among the Tier 2 proteins, TMEM106B had the broadest supporting evidence, including *cis*-eQTL colocalization in TOPCHeF failing left ventricular myocardium and GTEx left ventricular tissue, together with MR support in two proteomic cohorts.

## Discussion

In this study, we used GWAS-by-subtraction to separate HF genetic susceptibility from the component shared with BMI and integrated proteomic, colocalization, tissue-regulatory, transcriptomic, and druggability evidence to prioritize candidate proteins. We identified nine proteins associated with BMI-subtracted HF, with CELSR2 and CSF3 showing the strongest statistical support and several others supported by cardiac or cross-cohort evidence. Previous studies have shown that HF is genetically heterogeneous and arises through multiple upstream pathways, including obesity-related mechanisms^4,5^. Our findings further demonstrate that therapeutic target discovery can be performed on a biologically partitioned HF phenotype, thereby revealing pathways that may contribute to HF susceptibility beyond adiposity-related genetic effects.

The identification of LPA and the chromosome 1p13 CELSR2 locus links our findings to well-established lipid and vascular pathways. Previous genetic analyses suggested that the association between LPA and HF is largely attributable to coronary artery disease^4^, supporting an effect through atherosclerotic antecedents rather than a primary myocardial mechanism. Its persistence in BMI-subtracted overall HF is therefore consistent with a lipid-mediated pathway that is distinct from the genetic component shared with BMI. The CELSR2 finding requires additional consideration because functional studies at the 1p13 locus have historically emphasized hepatic SORT1 regulation and lipoprotein metabolism^41^. Although our plasma *cis*-pQTL MR and colocalization analyses specifically prioritize CELSR2 abundance, they do not definitively resolve the effector gene or exclude contributions from neighboring genes at this complex locus.

CSF3 points to a potentially important hematopoietic contribution to niHF. CSF3 encodes granulocyte colony-stimulating factor, a cytokine that regulates granulopoiesis and the mobilization of hematopoietic progenitor cells^42^. The risk-increasing association with niHF was among the strongest in our analysis, yet CSF3 lacked cardiac cell-type expression and cardiac eQTL colocalization support. This pattern raises the possibility that its association operates through circulating immune cells or another extracardiac pathway. However, lifelong genetically predicted differences in circulating CSF3 abundance should not be assumed to reproduce the effects of short-term recombinant G-CSF administration, and the relevant cellular mechanism remains to be established.

TMEM106B and DAG1 showed more direct connections to myocardial biology. TMEM106B is a lysosomal membrane protein involved in lysosomal morphology, trafficking, and function^43^. Its association was supported across proteomic cohorts, and the plasma *cis*-pQTL colocalized with TMEM106B expression in both GTEx left ventricle and TOPCHeF failing myocardium. Together with its cardiomyocyte expression, these findings raise the possibility that altered lysosomal homeostasis contributes to HF susceptibility, although the cardiac functions of TMEM106B remain largely unexplored. DAG1 encodes dystroglycan, a component of the dystrophin-associated glycoprotein complex that links the extracellular matrix to the intracellular cytoskeleton^44^. Its cardiomyocyte enrichment marked downregulation in failing myocardium, and association of higher genetically predicted abundance with lower HF risk provide complementary evidence implicating maintenance of cardiomyocyte structural integrity.

RSPO3 and GPX1 illustrate the importance of biological context when interpreting the direction of MR associations. RSPO3 is a secreted regulator of Wnt signaling with established roles in vascular and stromal biology^45^. Its fibroblast expression and inverse association with HF suggest a possible role in cardiac remodeling, but its upregulation in failing myocardium was directionally discordant with the MR result and may reflect a compensatory response. GPX1 encodes a major peroxide detoxifying enzyme and is generally considered protective against oxidative injury^46^. The association of higher genetically predicted GPX1 abundance with greater HF risk is therefore not readily explained by a simple antioxidant mechanism and may reflect differences between circulating abundance and intracellular enzymatic activity.

The therapeutic implications are protein specific. The association of higher LPA abundance with greater HF risk is directionally consistent with Lp(a)-lowering strategies and provides a clinically recognizable positive control^47,48^. In contrast, the RSPO3 and CSF3 findings suggest that human genetics may also provide insight into potential on-target cardiovascular effects of therapeutic modulation. Because higher genetically predicted abundance of these proteins was associated with lower HF risk, inhibition of their activity could theoretically have unintended cardiovascular consequences. Nevertheless, the effects of lifelong genetically predicted differences in protein abundance may differ substantially from those of pharmacological intervention. Further mechanistic and experimental studies will be needed to clarify the clinical implications of these observations.

This study has several strengths. We combined GWAS-by-subtraction with proteome-wide *cis*-pQTL MR and colocalization analyses to identify associations with HF beyond the genetic component shared with BMI. The use of four independent proteomic cohorts spanning SomaScan and Olink platforms enabled assessment of cross-cohort and cross-platform consistency. We further integrated tissue eQTL colocalization, cardiac cell-type expression, myocardial differential expression, and druggability to place the genetic findings in biological and therapeutic context. Finally, the recovery of established cardiovascular signals, including LPA and CELSR2, supported the validity of our analytic approach.

Several limitations warrant consideration. First, GWAS-by-subtraction partitions the genetic component shared between BMI and HF but does not fully eliminate all adiposity-related biology; residual genetic correlation with BMI may still remain. Second, most MR analyses relied on single *cis*-pQTL instruments, limiting pleiotropy-robust sensitivity analyses. Third, plasma *cis*-pQTLs reflect regulation of circulating protein abundance and may not capture protein regulation or function within the myocardium. The GTEx, TOPCHeF, and ReHeaT2 analyses provide corroborating tissue and cell-type context but do not definitively establish the relevant tissue, cell type, or mechanism. Similarly, concordance between MR associations and differential myocardial expression should be interpreted cautiously because expression changes in failing hearts may represent consequences of disease, treatment effects, or compensatory responses. Fourth, the ejection-fraction subtype analyses were underpowered, particularly for niHFpEF. Replication in larger subtype-resolved HF GWAS, independent proteomic cohorts, and additional cardiac molecular datasets will be needed to validate these findings and clarify their biological and therapeutic relevance.

In conclusion, integrating GWAS-by-subtraction with proteome-wide MR and colocalization identified nine circulating proteins associated with HF genetic susceptibility beyond the component shared with BMI. These findings provide new insight into the biology of HF not fully explained by adiposity and may inform future efforts to refine disease mechanisms, identify therapeutic targets, and develop more biologically grounded approaches to HF prevention and treatment.

## Nonstandard Abbreviations and Acronyms

BMI: body mass index
eQTL: expression quantitative trait locus
GWAS: genome-wide association study
HERMES: Heart Failure Molecular Epidemiology for Therapeutic Targets (consortium)
HF: heart failure
niHF: nonischemic heart failure
niHFpEF: nonischemic heart failure with preserved ejection fraction
niHFrEF: nonischemic heart failure with reduced ejection fraction
LD: linkage disequilibrium
LVEF: left ventricular ejection fraction
MR: Mendelian randomization
PP.H4: posterior probability of a shared causal variant
pQTL: protein quantitative trait locus
UKB-PPP: UK Biobank Pharma Proteomics Project

## Declarations

### Ethics approval and consent to participate

This research was conducted using secondary data and has been determined to be exempt from IRB review under the University of Wisconsin-Madison Institutional Review Board policies with a formal IRB waiver. All summary level data are publicly available.

### Data and resources

Analyses were performed in R v4.5.2 (https://www.r-project.org/). We used GenomicSEM (https://github.com/GenomicSEM/GenomicSEM), UCSC liftOver, PLINK v1.9 (http://pngu.mgh.harvard.edu/purcell/plink/), TwoSampleMR v0.7.5 (https://mrcieu.github.io/TwoSampleMR/), PWCoCo (https://github.com/jwr-git/pwcoco), SharePro v5.0 (https://github.com/zhwm/SharePro_coloc).

## Acknowledgments

C.-Y.S. acknowledges the Social Sciences Computing Core at the University of Wisconsin-Madison for providing computing resources for conducting these analyses.

## Author Contributions

C.-Y.S. conceived and designed the study, performed the analyses, interpreted the results, and drafted the manuscript. C.-Y.S. and T.L. revised the manuscript for important intellectual content and approved the final version.

## Supplementary Figure Legends

**Supplementary Figure 1.**
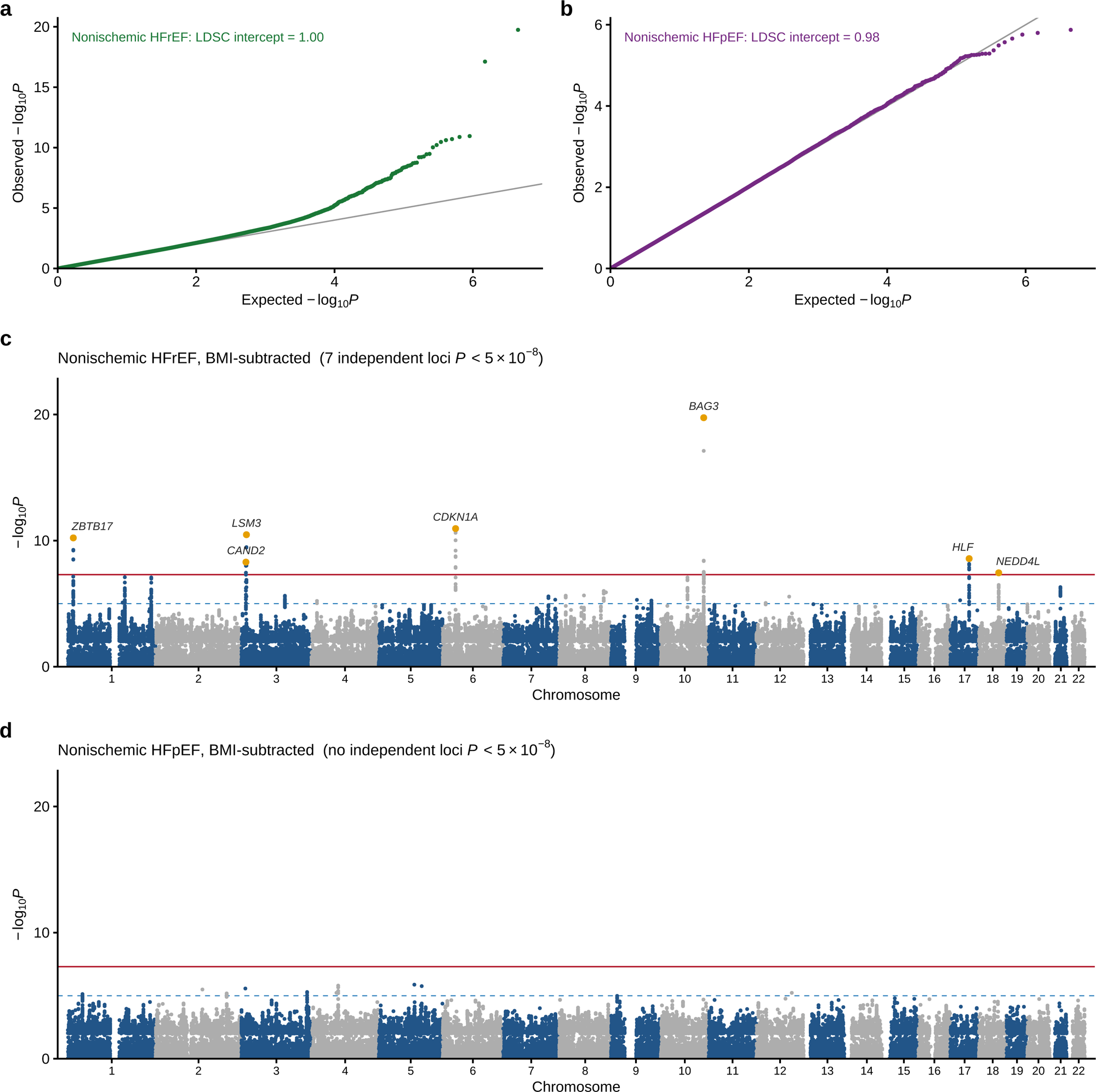
BMI-subtracted genome-wide association studies of lower-powered nonischemic heart failure subtypes. Quantile-quantile plots (a and b) and Manhattan plots (c and d) for the BMI-subtracted nonischemic heart failure with reduced ejection fraction (niHFrEF; a and c) and nonischemic heart failure with preserved ejection fraction (niHFpEF; b and d) genome-wide association studies. The quantile-quantile plots show the linkage disequilibrium score regression intercept. In the Manhattan plots, the red horizontal line indicates genome-wide significance (*P* = 5×10^-8^), and the blue dashed line indicates the suggestive threshold (*P* = 1×10^-5^). Lead variants at genome-wide significant loci are labeled with the nearest protein-coding gene, as in Figure 1. The complete locus list is provided in **Supplementary Table 3**. The BMI-subtracted niHFrEF GWAS identified 7 genomw-wide significant independent loci, whereas the niHFpEF GWAS, which included 3,590 cases, identified none.

**Supplementary Figure 2.**
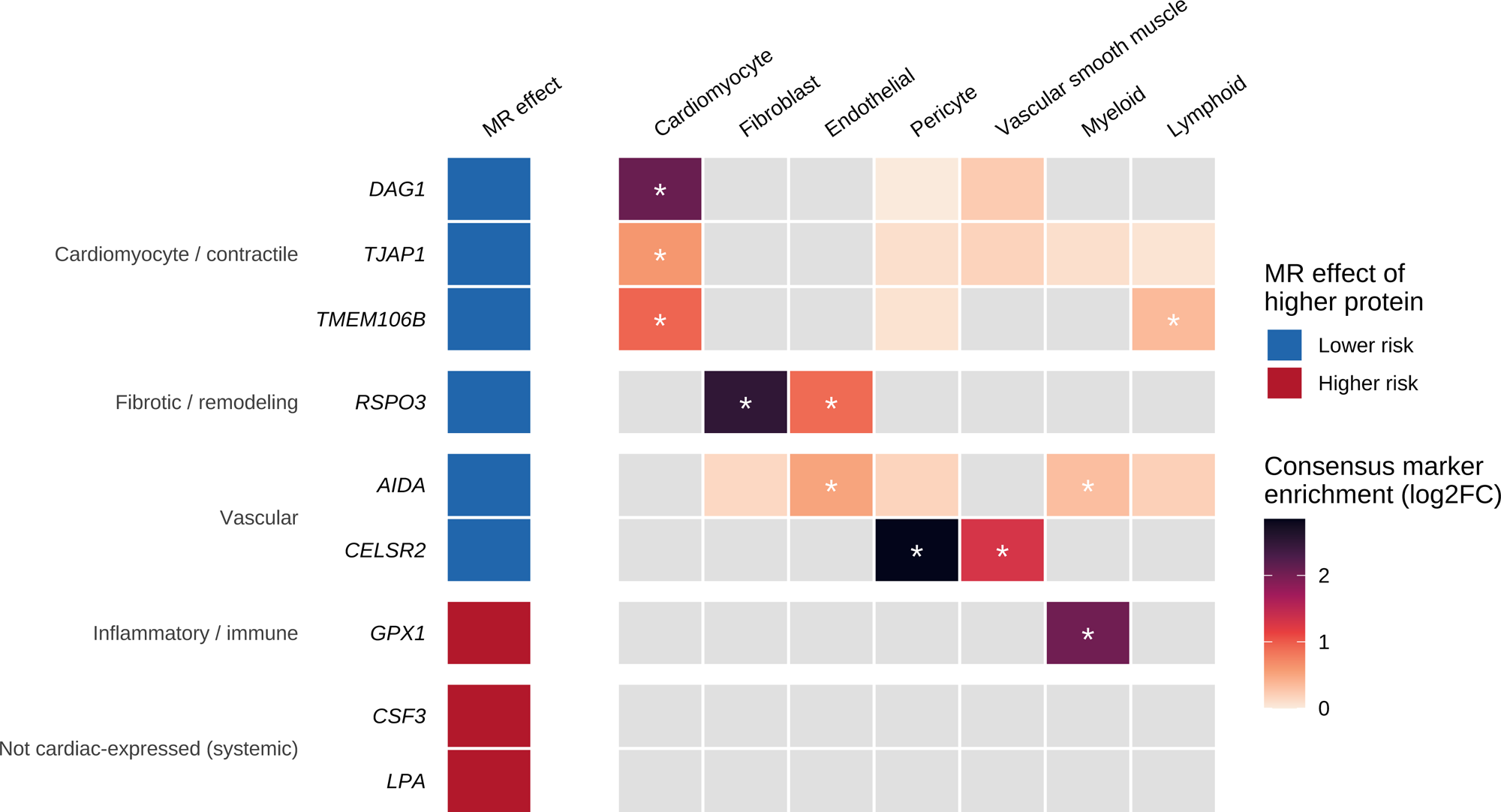
Cardiac cell-type expression of prioritized proteins in ReHeaT2. Heatmap showing the cardiac cell-type expression patterns of the nine prioritized proteins across seven cell types from the ReHeaT2 single-nucleus integration. Rows are grouped by broad biological category. Tile shading represents consensus marker enrichment for each gene, expressed as the mean log_2_ fold change relative to the remaining cell types; darker shading indicates greater enrichment. Asterisks indicate adjusted Fisher exact test *P* < 0.05. Gray tiles indicate that the gene was not identified as a consensus marker for the corresponding cell type. Fully gray rows indicate proteins without cardiac cell-type marker evidence in ReHeaT2. The left annotation strip shows the direction of the Mendelian randomization association for higher genetically predicted protein abundance with BMI-subtracted HF, with blue indicating lower HF risk and red indicating higher HF risk.

